# Balanced Electrolyte solution versus Saline Trial for Diabetic KetoAcidosis-Statistical Analysis Plan

**DOI:** 10.1101/2025.10.26.25337451

**Authors:** Anthony Devaux, Qiang Li, Laurent Billot, Bala Venkatesh, Mahesh Ramanan BEST-DKA Management Committee

## Abstract

**Introduction:** Fluid therapy is a cornerstone of acute management of diabetic ketoacidosis (DKA), yet the optimal fluid type is unknown. The Balanced Electrolyte Solution versus Saline Trial for Diabetic Ketoacidosis (BEST-DKA) trial has been designed to test the hypothesis that for patients with moderate to severe DKA, fluid therapy with Plasma-Lyte 148, compared with 0.9% saline, will lead to increased days alive and out of hospital to day-28 after enrolment.

**Purpose:** To describe the statistical analysis plan for the BEST-DKA trial. Design, settings and participants: BEST-DKA is a currently recruiting cluster-randomised, crossover randomised controlled trial which will recruit a minimum of 400 patients with moderate or severe DKA at 22 hospitals in Australia, which will provide >91.4% power to detect a change of 2 days in the primary outcome, days alive and out of hospital to day-28 after enrolment. Hospitals will be randomised to either blinded saline or Plasma-Lyte 148 for a 12-month period, and then crossover to the alternate fluid for the next 12-month period.

**Conclusion:** BEST-DKA started enrolment in March 2024 and recruitment is on target with scheduled completion in July 2026.

**Administrative information:** *Study identifiers:*

- ClinicalTrials.gov register Identifier: NCT05752279.

*Revision history:* 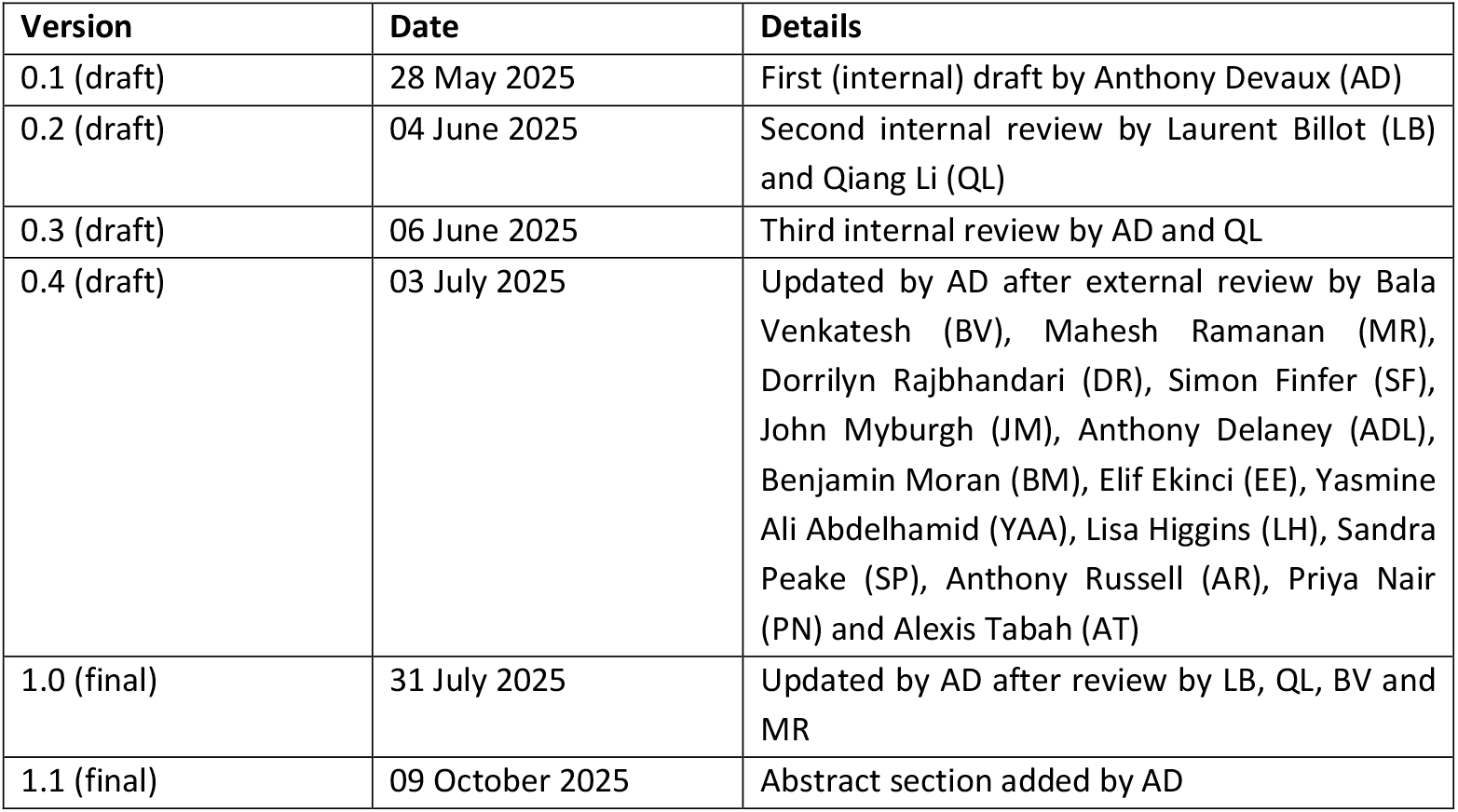

*Contributors to the statistical analysis plan:* 1.3.1
Roles and responsibilities

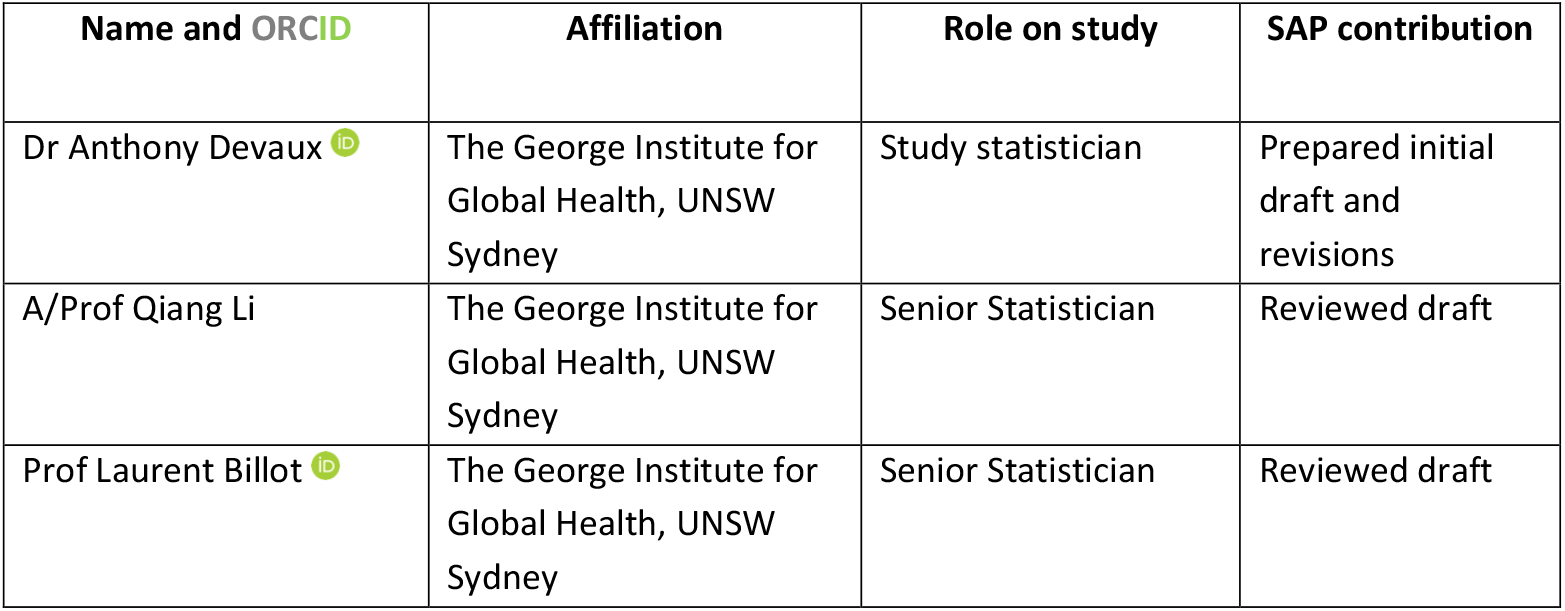
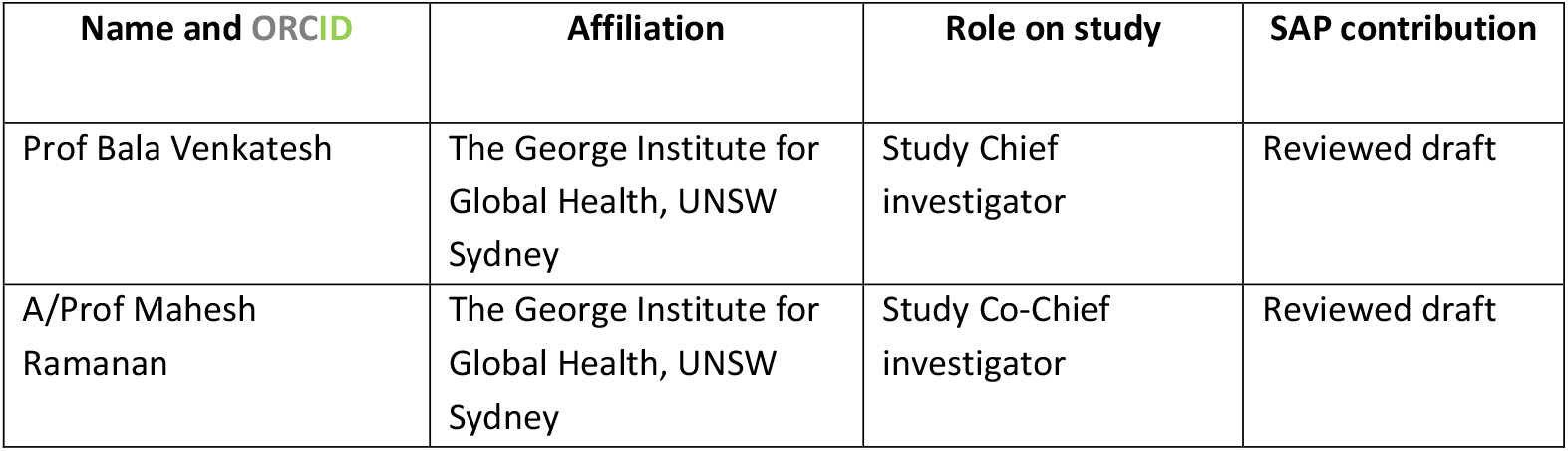

1.3 Contributors to the statistical analysis plan 1.3.1 Roles and responsibilities
| Name and ORCID | Affiliation | Role on study | SAP contribution |
| --- | --- | --- | --- |
| Dr Anthony Devaux | The George Institute for Global Health, UNSW Sydney | Study statistician | Prepared initial draft and revisions |
| A/Prof Qiang Li | The George Institute for Global Health, UNSW Sydney | Senior Statistician | Reviewed draft |
| Prof Laurent Billot | The George Institute for Global Health, UNSW Sydney | Senior Statistician | Reviewed draft |

| **Name and ORCID** | **Affiliation** | **Role on study** | **SAP contribution** |
| --- | --- | --- | --- |
| Prof Bala Venkatesh | The George Institute for Global Health, UNSW Sydney | Study Chief investigator | Reviewed draft |
| A/Prof Mahesh Ramanan | The George Institute for Global Health, UNSW Sydney | Study Co-Chief investigator | Reviewed draft |

1.3.2
Approvals
The undersigned have reviewed this plan and approve it as final. They find it to be consistent with the requirements of the protocol as it applies to their respective areas. They also find it to be compliant with ICH-E9 principles and in particular confirm that this analysis plan was developed in a completely blinded manner, i.e. without knowledge of the effect of the intervention(s) being assessed.

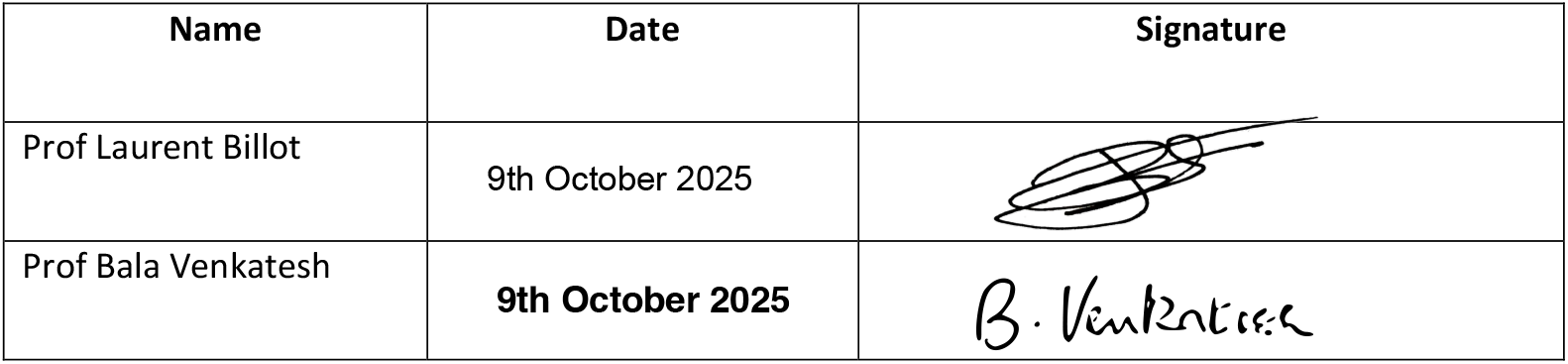

| **Name** | **Date** | **Signature** |
| --- | --- | --- |
| Prof Laurent Billot | 9th October 2025 |  |
| Prof Bala Venkatesh | **9th October 2025** |  |

## 2 Introduction

### 2.1 Study synopsis

Diabetic ketoacidosis (DKA) is a life-threatening complication of diabetes mellitus, described in patients with both type-1 diabetes, and type-2 diabetes. BEST-DKA is a Phase 3 cluster-crossover, blinded, pragmatic, randomised, controlled trial comparing the effects of saline or buffered crystalloid solution in patients with moderate to severe DKA treated in the Emergency Department (ED) and/or Intensive Care Unit (ICU) at twenty hospitals in Australia. Each hospital will be randomised to use either 0.9% saline or buffered crystalloid solution (Plasma-Lyte® 148) for a period of 12 months before crossing over to the alternate fluid for the next 12 months. The blinded study fluid will be used for all resuscitation and maintenance purposes for included patients.

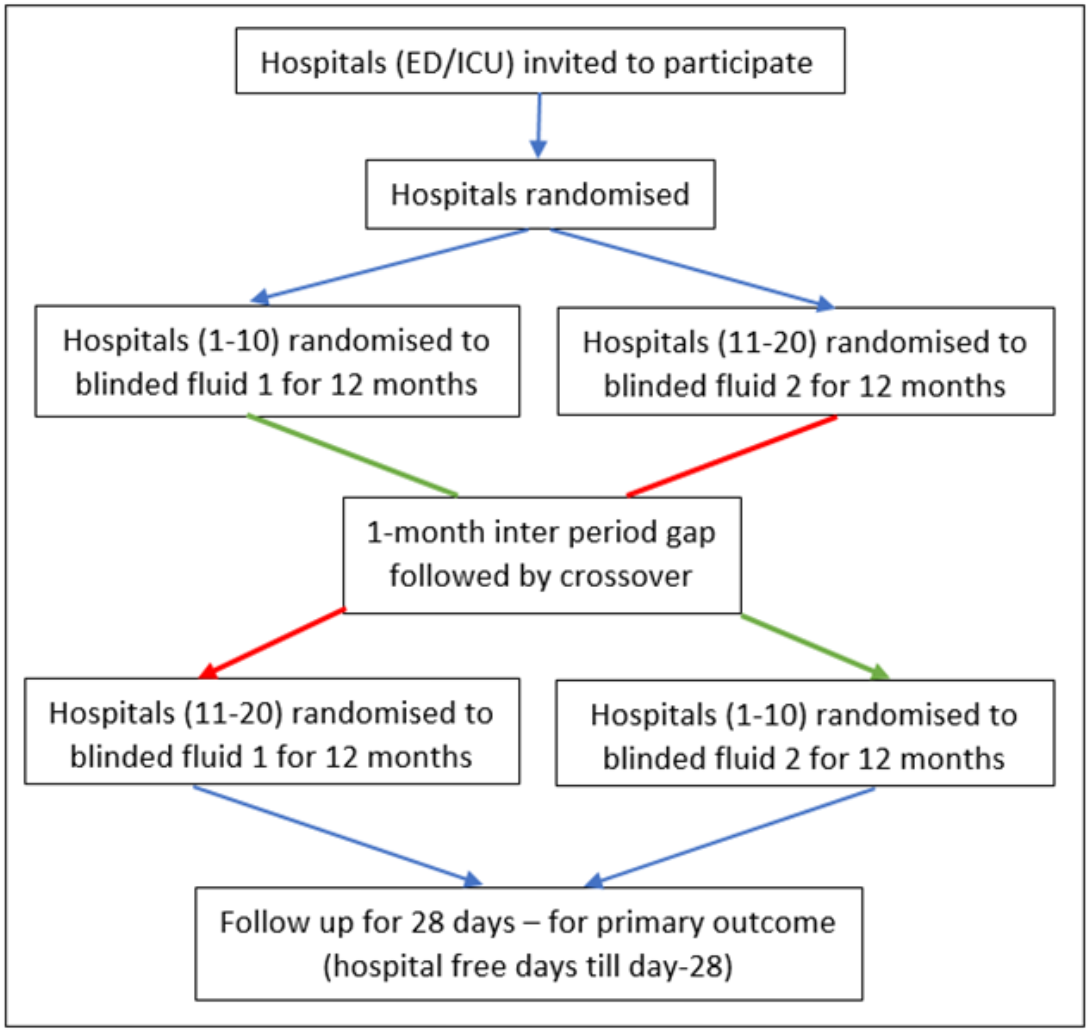

### 2.2 Study population

#### 2.2.1 Site eligibility criteria

- **Inclusion**
  - Hospitals with an ED and a history of treating at least 20 DKA patients who are transferred to the ICU per year as previously recorded in the ANZICS CORE adult patient database
  - Clinical management of DKA is in accordance with hospital or national guidelines
- **Exclusions**
  - Unwilling or unable to follow trial protocols
  - Unable to capture the minimum data set required for the study

#### 2.2.2 Patient eligibility criteria

- **Inclusion**
  - Patients in the ED with a primary diagnosis of DKA for whom both 0.9% saline and Plasma-Lyte® 148 are considered appropriate fluids
  - Blood glucose level > 14mmol/L
  - pH < 7.25
  - Serum bicarbonate <15 mmol/L
  - Elevated anion gap > 12mEq/L
  - Ketones positive on finger-prick measurement
  - In the judgement of the treating clinician critical care area admission is required
- **Exclusions**
  - Age less than 18 years
  - Patients who have received more than 2000ml of non-study fluid prior to study enrolment
  - Serum Na > 155 or <120 mmol/L
  - Contraindication to either study fluid e.g. previous allergic reaction to Plasma-Lyte® 148
  - Patients with hyperosmotic hyperglycaemic non-ketotic syndrome
  - Other clinical conditions that preclude large volumes of fluid resuscitation
  - Previous inclusion in BEST-DKA trial study within the last 28 days

Patients readmitted to the ICU within 28 days will be re-enrolled into the study and receive study interventions if they meet inclusion criteria and do not have any exclusion criteria. They will be counted as the same enrolment for study analysis.

### 2.3 Interventions

Sites will be randomly allocated to use either Plasma-Lyte® 148 or 0.9% saline for all eligible patients with moderate to severe DKA during the first 12-month period and then crossover to the alternate fluid for the second 12-month period. BEST-DKA study fluids will be started whilst the participant is in the emergency department (ED). The study treatments will be supplied in identical 1000 mL bags with standardised fluid administration sets without revealing the fluid type. Both fluids are colourless, clear solutions and macroscopically indistinguishable. Both participants and study investigators will be blinded to study treatment allocation. Study fluids will be used for all resuscitation and maintenance purposes. Other crystalloid fluids may be used as carrier fluids for the infusion of any drug for which either Plasma-Lyte® 148® or 0.9% saline is considered incompatible; in such instances 5% glucose should be used whenever possible to minimise exposure to open-label Plasma-Lyte® 148® and 0.9% saline.

#### 2.3.1 Intervention arm

In ICUs allocated to the intervention arm, patients will receive Plasma-Lyte® 148 fluid therapy.

#### 2.3.2 Control arm

In ICUs allocated to the control arm, patients will receive 0.9% saline fluid therapy.

### 2.4 Outcomes

#### 2.4.1 Primary outcome

- Days alive and out of hospital up to 28 days after study enrolment

The primary outcome is calculated as 28 minus total number of days spent as an inpatient in any hospital during the first 28 days from the date of study enrolment. This includes the index hospital admission and any hospital readmissions during the 28-day follow-up period. Any part of a day spent as an inpatient in a hospital will count as one day. Any patient who dies within the 28-day period will be assigned a score of zero.

Examples of this calculation are provided below:

- A patient who survived and is discharged from hospital on day 4 after enrolment into trial is assigned a value of 24.
- A patient who survived and is discharged from hospital on day 4 after enrolment into trial and readmitted on Day 8 for another 4 days is assigned a value of 20.

#### 2.4.2 Secondary outcomes

- Days alive and out of ICU up to 28 days after study enrolment
- Number of ICU readmissions up to 28 days after study enrolment
- Number of hospital readmissions up to 28 days after study enrolment
- Proportion of patients in each stage of Acute Kidney Injury using KDIGO criteria ^1^up to 72 hours in a critical care area, where the stages are defined as followed:
  - Stage 1: increase of serum creatinine ≥ 1.5 and < 2.0 from baseline
  - Stage 2: increase of serum creatinine ≥ 2.0 and < 3.0 from baseline
  - Stage 3: increase of serum creatinine ≥ 3.0 from baseline
- Number of episodes of post-randomisation decrease in Glasgow coma score (GCS) by more than 2 in the first 24 hours
- Time to resolution (< 1 mmol/L) of ketosis (in hours)
- Cumulative insulin dosage in the first 48 hours
- Duration of intravenous insulin infusion in the ICU
- Cumulative potassium replacement in the first 48 hours
- Quality of life 28 days after study enrolment as measured by the EuroQoL 5D-5L ^2^ instrument
- Modified Fatigue score ^3^ 28 days after study enrolment

#### 2.4.3 Process measures

- Serum acetoacetate and β-hydroxybutyrate at 12, 24 and 48 hours
- Serum base excess at 6, 12 and 24 hours
- Serum sodium, potassium and chloride concentration at 24 hours

#### 2.4.4 Tertiary outcomes

- Cost-effectiveness analysis

#### 2.4.5 Safety outcomes

- Number of episodes of Hypoglycaemia
- Number of episodes of Hypophosphataemia
- Number of episodes of Hypernatraemia
- Number of Episodes of Glasgow Coma Score decrease by 2 or more points

### 2.5 Randomisation and blinding

ICUs were randomised in the pre-trial period to manage included patients with either Plasma-Lyte® 148 as fluid therapy in the first 12-month period and then 0.9% saline in the second 12-month period, or 0.9% saline in the first period and Plasma-Lyte® 148 in the second period. The fluids will be delivered as “Fluid A” or “Fluid B” to be stored and used for the entire first study period with crossover to the alternate fluid for the second study period. Study participants, treating clinicians, study investigators and data collectors will be blinded to study treatment allocation.

### 2.6 Statistical hypotheses

The primary statistical hypotheses are as follows:

- **Null hypothesis**: no difference in the days alive and out of hospital up to 28 days between patients randomised to Plasma-Lyte® 148 (µ_1_) and 0.9% saline (µ_0_) i.e. µ_1_ = µ_0_
- **Alternative hypothesis (2-sided)**: µ_1_ ≠ µ_0_, those randomly assigned to receive Plasma-Lyte® 148 will experience more days alive and out of hospital to day 28 compared to those randomly assigned to receive saline.

### 2.7 Sample size

BEST-DKA initially aimed to recruit patients from 20 hospitals each recruiting an expected 10-12 patients per period (i.e. 20-24 patients per hospital), generating an anticipated sample size range between 400-480 patients. Due to lower recruitment rate than expected in some hospital, two hospitals were added bringing the total to 22 hospitals.

Assuming an mean of 21.4 days and SD = 6.46 in 0.9% saline group, and mean of 23.3 days and SD = 3.86 in Plasma-Lyte 148 group, an exchangeable correlation structure with an intra-cluster correlation coefficient (ICC) of 0.01 and a potential withdrawal and loss to follow-up rate of 1% (based on Phase II data from the SCOPE-DKA trial ^4^), the recruitment of 400 and 480 participants provides 91.4% and 94.0% power respectively to identify a difference between arms of 2.0 days alive and out of hospital, with a 2-sided α = 0.05.

## 3 Statistical analysis

### 3.1 Statistical principles

#### 3.1.1 Interim analysis and stopping rules

One interim analysis will be conducted when all sites have finished recruitment and follow-up until 28 days over the first 12-month intervention period. Kim and deMets alpha and beta spending functions have been used to define the stopping rules for efficacy/harm and futility (see Table 1 below).

**Table 1.** Stopping rules.

| Analysis stage | Interim | Final |  |
| --- | --- | --- | --- |
| Planned information rate | 50% | 100% |  |
| Cumulative alpha spent | 0.35% | 5.00% |  |
| Cumulative beta spent | 3.66% | 10.00% |  |
| Cumulative power | 28.90% | 90.00% |  |
| Significance threshold (two-sided) | 0.0035 | 0.0488 |  |
| Efficacy boundary (z-value scale) | 2.921 | 1.97 |  |
| Futility boundary (z-value scale) | 0.592 |  |  |
| *Lower efficacy boundary (t)* | *-2.505* | *-1.179* |  |
| *Upper efficacy boundary (t)* | *2.505* | *1.179* |  |
| *Upper futility boundary (t)* | *0.5* |  |  |
| Exit probability for efficacy (under H0) | 0.35% | 4.65% | $\alpha$ |
| Exit probability for futility (under H0) | 44.61% | 50.39% |  |
| Overall exit probability (under H0) | 44.96% | 55.04% |  |
| Exit probability for efficacy (under H1) | 28.90% | 61.10% | *power* |
| Exit probability for futility (under H1) | 3.66% | 6.34% | $\beta$ |
| Overall exit probability (under H1) | 32.56% | 67.44% |  |

**Figure 1:**
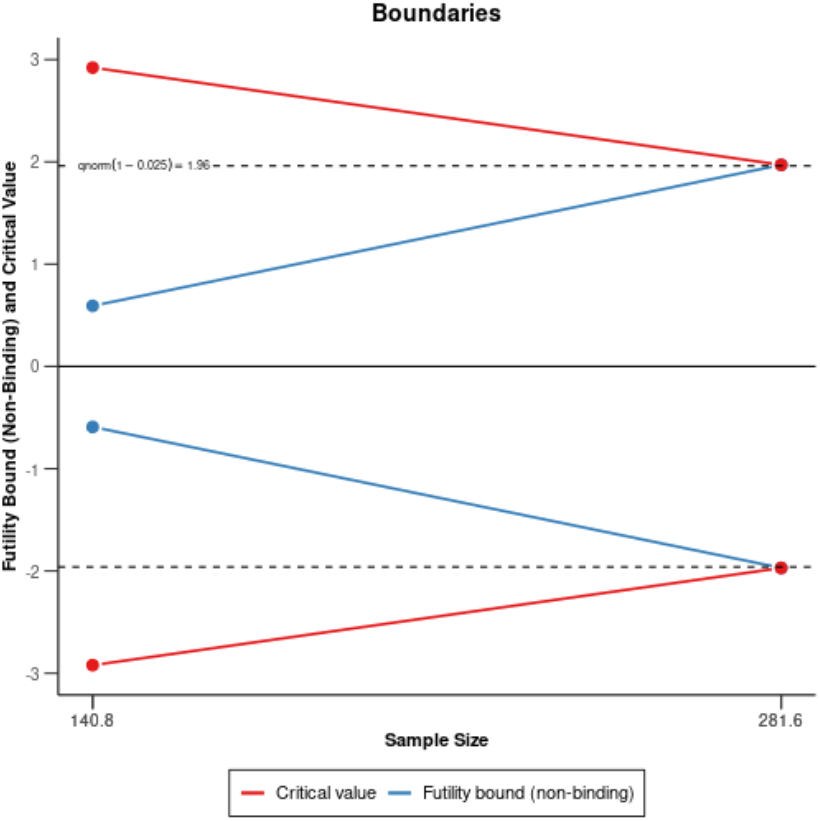
Boundaries for efficacy/harm and futility.

The trial would be stopped at the interim for:

- <u>Efficacy</u>: if the z statistic is higher than 2.921. Assuming an individually randomised trial with a SD of 5, this corresponds to a difference in DAOH of more than 2.5 days
- <u>Harm</u>: if the z statistic is lower than −2.921 Assuming an individually randomised trial with a SD of 5, this corresponds to a difference in DAOH of less than −2.5 days
- <u>Futility</u>: if the z statistics is between 0.592 and −0.592. Assuming an individually randomised trial with a SD of 5, this corresponds to a difference in DAOH comprised between −0.5 day and +0.5 day

A non-binding futility boundary has been set if the treatment effect falls between −0.5 and 0.5 days at the interim analysis. A recommendation to stop for futility will be non-binding, and at the discretion of the BEST-DKA management committee as to whether recruitment will be ceased or not.

If deemed appropriate the DSMC can recommend the BEST-DKA Study Management Committee should:

- terminate the study early if there is clear and substantial evidence of benefit;
- terminate the study early if the data suggests the risk of adverse events substantially outweighs the potential benefits; or
- recommend termination of the study early for futility.

#### 3.1.2 Final analysis

Final analyses of the primary outcome including sensitivity analyses will all be conducted using a two-sided significance level of 4.88%. For the secondary clinical outcomes, no formal adjustment for multiple comparisons will be applied. To reduce the risk of over-interpretation of results due to multiplicity, p-values will not be presented. Instead, emphasis will be placed on the magnitude and precision of effect estimates, including confidence intervals, to guide interpretation.

#### 3.1.3 Statistical software

Analyses will be conducted primarily using R software (version 4.5 or above).

### 3.2 Analysis populations

Due to the cluster-crossover design, the group allocation for a patient is determined by the site and by the period during which they participated, regardless of treatment adherence. The intention-to-treat analysis set will be used to assess both effectiveness and safety. The flow of patients through the study will be displayed in a CONSORT diagram.

### 3.3 Analysis principles

- Analyses will be conducted on an intention-to-treat (ITT) basis – i.e., analysing all patients according to the group to which they were assigned regardless of treatment compliance.
- The primary outcome will be analysed using a two-sided significance level α = 4.88%.
- The secondary outcomes will be analysed using a two-sided significance level α = 5%, before correction for multiplicity testing using Holm-Bonferroni method.
- This analysis plan, and the primary manuscript, will only include analyses up to 28 days following enrolment.
- Quality of life and fatigue scores at 28 days post enrolment will be presented in a separate publication.
- Pre-specified subgroup analyses will be conducted regardless of whether statistically significant treatment effect on the primary outcome is observed in the overall sample.
- Continuous variables will be analysed using standard parametric methods (e.g. t-test).
- Tests of normality will not be conducted.

### 3.4 Baseline analyses

Description of the baseline characteristics will be presented by treatment group (see Table 1). Discrete variables will be summarised by frequencies and percentages. Percentages will be calculated according to the number of patients for whom data are available and the number of missing data reported. Continuous variables will be summarised by using mean and SD, and median and interquartile range (Q1-Q3).

### 3.5 Treatment compliance

Compliance with the randomized intervention will be summarised using the following variables:

- Time on study treatment defined as the number of days between the first and last study fluid administration (up to 28 days)
- Cumulative dose of study drug received (mL) up to 28 days
- Number and proportion of patients who received 500mL or more of the study fluid (Plasma-Lyte® 148 or 0.9% saline) as an open label fluid in the ICU (post-randomisation) when they were assigned to the other fluid
- Number and proportion of patients for whom study treatment was permanently discontinued whilst in the ICU together with the main discontinuation reason

Time on study treatment and cumulative dose, will be summarised using means, SD, median and quartiles with differences between treatment groups tested using a linear model with a random site effect. Differences in proportions will be compared using a logistic model with a random site effect.

### 3.6 Protocol deviations

Protocol deviations will be categorised and reported as the number and proportion of subjects experiencing a deviation among the following:

- Enrolment of an ineligible patient.
- Non-study fluid administered (Saline (any concentration), Plasma-Lyte, Hartmann’s, Glucose solution (any concentration), Mixed glucose/saline solution (e.g. 4% glucose/0.18% NaCl), Gelatin based fluid, Starch based fluid, Other fluid)
- Other

### 3.7 Analysis of the primary outcome

The primary outcome is the days alive and out of hospital up to 28 days after enrolment. The primary intervention effect will be estimated as the mean difference (MD) of days alive and out of hospital between Plasma-Lyte® 148 and 0.9% saline obtained from a linear mixed model (defined below).

#### 3.7.1 Interim analysis

The single interim analysis will be performed after the first 12-month intervention period, and will only include data collected during the first period. The primary outcome of days alive and out of hospital up to 28 days will be modelled using a hierarchical linear model as follows:

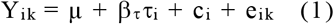

where:

Y_ik_ denotes the days alive and out of hospital of subject k (k=1,…,m_i_) within cluster I (i=1,…, N) where m_i_ indicates the number of subjects within cluster i

µ is the overall intercept

*τ*_i_ is a dummy variable indicating group allocation for cluster i with *τ*_i_ = 1 for intervention (Plasma-Lyte® 148) and *τ*_i_ = 0 for control (0.9% saline)

β_*τ*_ is the parameter of interest estimating the fixed effect of the Plasma-Lyte® 148 intervention

c_i_ is a random cluster effect with c_i_ ∼ N(0, *σ*_c_ ^2^)

e_ik_ are individual-level errors following a Gaussian distribution with e_i_ ∼ N(0, *σ*_e_ ^2^)

The effect of the intervention β_*τ*_ will be presented as the mean difference (MD) of days alive and out of hospital and its 95% confidence interval (CI) using the control arm as the reference (i.e. where an MD greater than 0 corresponds to an increase in days alive and out of hospital in the Plasma-Lyte® 148 intervention arm compared to the 0.9% saline control arm). The Kenward-Rodger [6] correction will be applied to estimate the number of degrees of freedom as it has been shown to improve estimation with a small number of clusters. The z-statistic which corresponds to the test of the intervention effect (H_0_: *β*_*τ*_ = 0) will be obtained from the random effect model. It will be compared to the boundaries on the z-value scale to assess whether the pre-specified efficacy or futility boundaries are crossed (Table 1, Section 3.1.1).

#### 3.7.2 Final analysis

For the final analysis, a fixed period effect and a random cluster-period effect will be added to model 1 to account for clustering of participants within sites as well as the cross-over design (see “random-random” model (equation 11) in Morgan et al ^5^). The corresponding model can be written as follows:

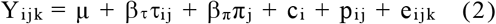

where:

Y_ij k_ denotes the days alive and out of hospital of subject k (k=1,…,m_ij_) in period j (j=1, 2) within cluster i (i=1,…, N) where m_ij_ indicates the number of subjects in period j within cluster i

µ is the overall intercept

*τ*_ij_ is a dummy variable indicating group allocation during period j for cluster i with *τ*_ij_ = 1 for intervention (Plasma-Lyte® 148) and *τ*_ij_ = 0 for control (0.9% saline)

*β*_*τ*_ is the parameter of interest estimating the fixed effect of the Plasma-Lyte® 148 intervention

π_j_ is a dummy variable indicating the period with with π_j_ = 1 for the second period and π_j_ = 0 for the first period

*β*_π_ is a fixed period effect

c_i_ is a random cluster effect with c_i_ ∼ N(0, *σ*_c_ ^2^)

p_ij_ is a random cluster-period effect with p_ij_ ∼ N(0, *σ*_p_ ^2^)

e_ij k_ are individual-level errors following a Gaussian distribution with e_ij_ ∼ N(0, *σ*_e_ ^2^)

The effect of the intervention *β*_*τ*_ will be presented using the same methodology as described in section 3.7.1. Based on this model, two intra-cluster correlation coefficients (ICC) will be estimated, the first one (ρ_c_) corresponding to the correlation between two outcomes from the same cluster-period, the second one (ρ_p_) corresponding to the additional correlation between two outcomes from the same cluster-period compared to two outcomes from the same cluster but different periods. The two ICCs, ρ_c_ and ρ_p_, will be estimated as follow:

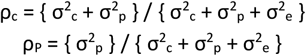

where:

*σ*^2^_c_ is the variance between the cluster means

*σ*^2^_p_ is the variance between the cluster-period means conditional on the cluster mean

*σ*^2^_e_ is the variance of the residual error.

The inter-period correlation (IPC), that is the correlation between two outcomes from the same cluster, but different periods will be estimated as:

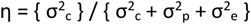

#### 3.7.3 Adjusted analyses

The linear mixed model described in Section 3.7.2 will be re-run after adjustment for the following individual baseline covariates:

- Age (continuous variable)
- Type of diabetes (categorical variable: Type 1, Type 2, Other)
- Anion gap at ED presentation (continuous variable)
- pH at ED presentation (continuous variable)

#### 3.7.4 Subgroup analyses

Prespecified subgroups analyses are planned for the primary outcome only, regardless of the statistical significance for the primary analysis:

- Type I vs. Type II diabetes
- Severity of acidosis (using first pH result in ED & last prior to enrolment and defined as pH of 7.00-7.25 or pH <7.0)
- Type of pre-enrolment fluid (Saline vs. buffered salt solutions)
- Above vs. below median of volume of open label pre-enrolment fluids
- Sex
- Patient on SGLT-2 inhibitors vs. those not on SGLT-2 inhibitors

Each subgroup analysis will be performed by adding the subgroup variable as well as its interaction with the intervention as fixed effects to the hierarchical linear model used for the primary analysis (see Section 3.7.2). Within each subgroup, summary measures will include mean and standard-deviation within each treatment arm, as well as the MD for treatment effect with 95% CI. The results will be displayed on a forest plot including the p-value for heterogeneity corresponding to the interaction term between the intervention and the subgroup variable.

#### 3.7.5 Treatment of missing data

The primary analysis will be performed on complete cases. If more than 5% of patients are excluded from this analysis due to missing data on days alive and out of hospital up to 28 days, we will further explore the impact of missing data by performing multiple imputations using fully conditional specification (FCS). The imputation model will include days alive and out of hospital up to 28 days, the randomised treatment arm, study site as well as baseline characteristics. Days alive and out of hospital up to 28 days will be imputed using a linear regression. One hundred sets of imputed data will be created and modelled to get 100 estimates, then pooled to obtain the mean difference and 95% confidence interval.

### 3.8 Analysis of secondary clinical outcomes

#### 3.8.1 Continuous outcomes

Continuous secondary clinical outcomes include days alive and out of ICU, number of ICU and hospital readmissions up to 28 days after enrolment, number of episodes of post-randomisation decrease in GCS by more than 2 in the first 24 hours, cumulative insulin dosage in the first 48 hours, duration of intravenous insulin infusion in the ICU and cumulative potassium replacement in the first 48 hours, EQ-5D-5L total score at 28 days and modified Fatigue score at 28 days. Those outcomes will be analyzed using the same approach as the primary outcome, described in section 3.7.2. The effect of the intervention will be reported as the mean difference and its 95% confidence interval.

#### 3.8.2 Categorical outcomes

The probability of hypokalaemia will be modeled using a hierarchical logistic regression using the same fixed and random effects as the primary outcome, described in section 3.7.2. The effect of the intervention will be reported as the odd-ratio and its 95% confidence interval.

The probability in each stage of Acute Kidney Injury defined by KDIGO criteria will be modelled using a hierarchical multinomial logistic regression using the same fixed and random effects as the primary outcome, described in section 3.7.2. The effect of the intervention will be reported for stage 2 vs. stage 1 and stage 3 vs. stage 1, as the odd-ratio and its 95% confidence interval.

#### 3.8.3 Time-to-event outcomes

The time to hospital discharge, time to ICU discharge and time to resolution of ketosis will be summarized using cumulative incidence functions. Medians and quartiles of time to discharge will be obtained from the cumulative incidence functions. The effect of the intervention will be estimated with a Cox model including the same fixed and random effects as the primary outcome, described in section 3.7.2. The effect of the intervention will be reported as the hazard-ratio and its 95% confidence interval.

### 3.9 Analysis of process measures

The process measures (serum concentration) will be presented by treatment group and overall (see supplementary Table 1), and will be summarised by using mean and SD, and median and interquartile range (Q1-Q3).

### 3.10 Analysis of tertiary outcome

Those analyses will be outlined in a separate health economic analysis plan.

### 3.11 Analysis of safety outcomes

The safety outcomes will be summarised as the number and proportion of patients experiencing at least one event. This will be done overall and by category. In addition, the total number of events will be reported. Proportions of patients with at least one SAEs and proportion of patients dead will be compared between treatment arms using Fisher’s exact test. A listing of all adverse events will be provided.

## Data Availability

This manuscript is a statistical analysis plan. It does not contain any data.

## 5 Main tables and figures

### 5.1 Tables

#### 5.1.1

**Table 1.**
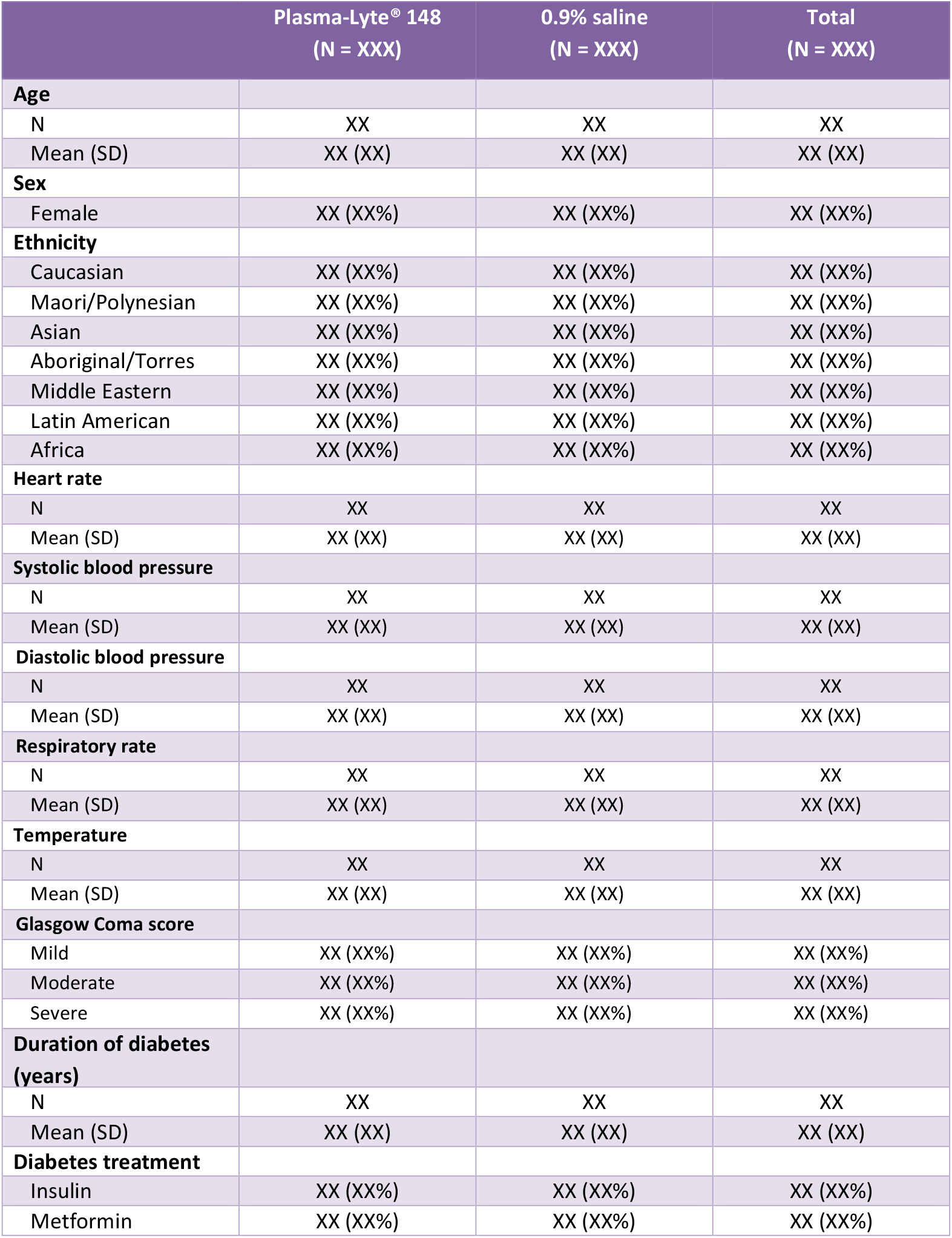
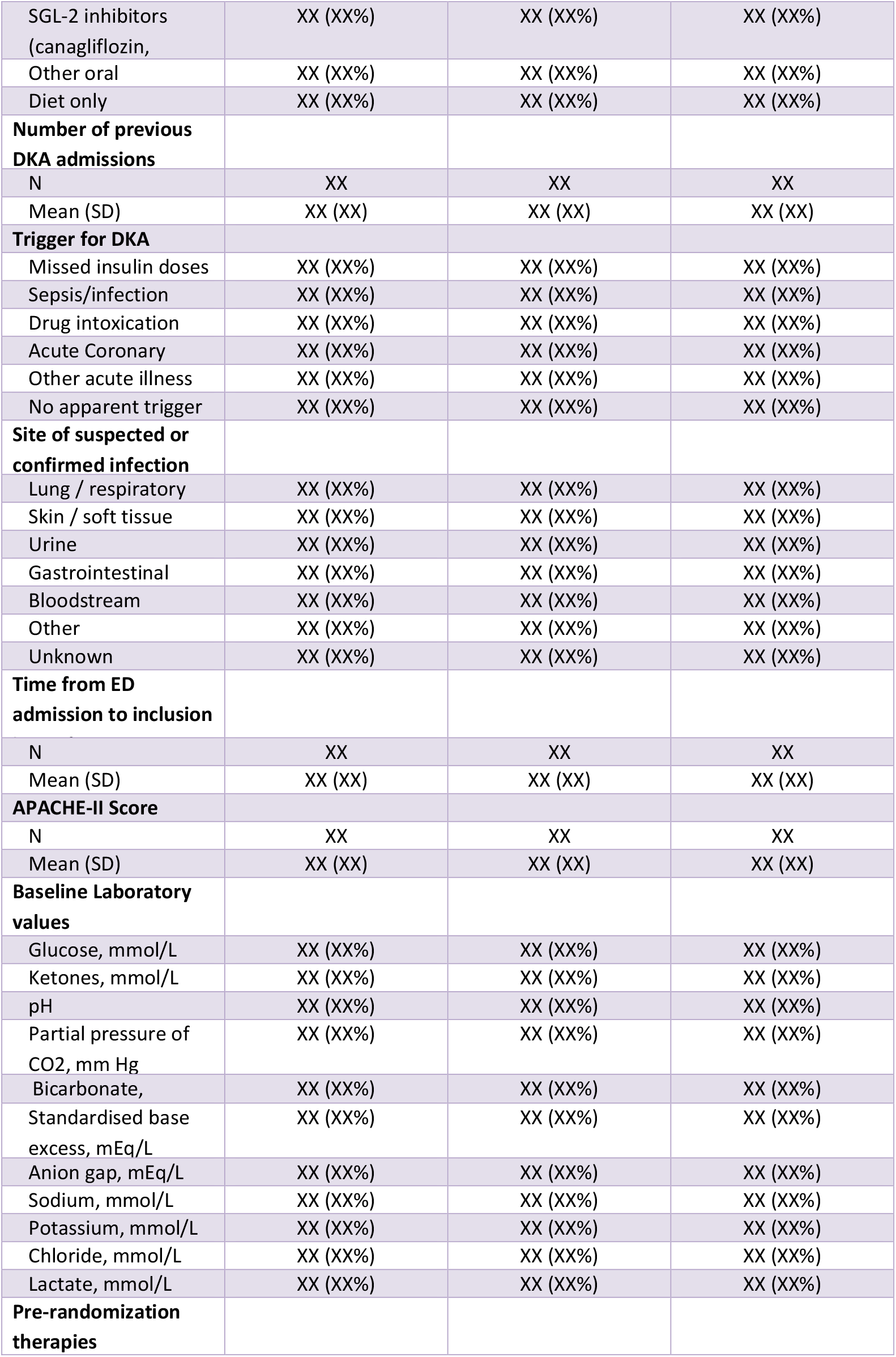
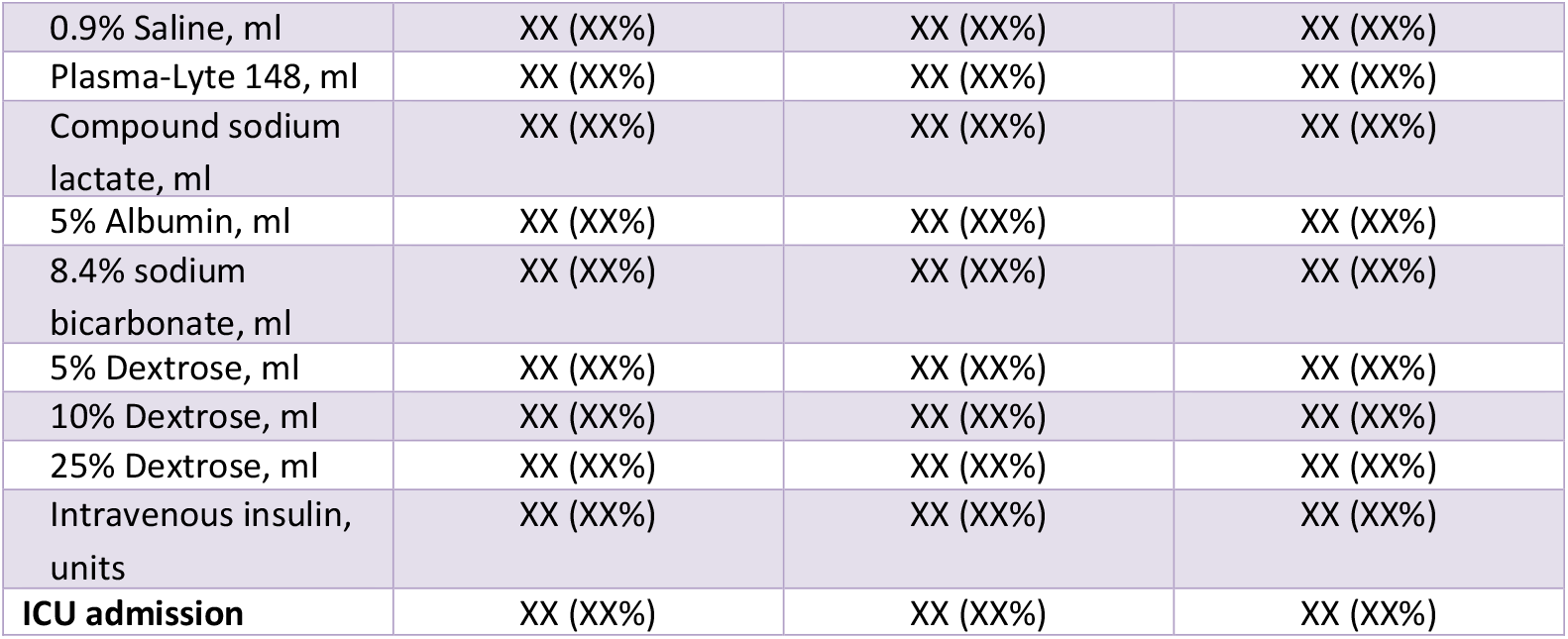
Baseline characteristics.

#### 5.1.2

**Table 2.** Treatments administered after inclusion.

|  | Plasma-Lyte®<br>148<br>(N = XXX) | 0.9% saline<br>(N = XXX) | Total<br>(N = XXX) |
| --- | --- | --- | --- |
| **Insulin administered up to 72 hours after inclusion in study** |  |  |  |
| N | XX | XX | XX |
| Mean (SD) | XX (XX) | XX (XX) | XX (XX) |
| **Fluids administered up to 72 hours after inclusion in study** |  |  |  |
| **BEST-DKA fluid** |  |  |  |
| N | XX | XX | XX |
| Mean (SD) | XX (XX) | XX (XX) | XX (XX) |
| **0.9% saline** |  |  |  |
| N | XX | XX | XX |
| Mean (SD) | XX (XX) | XX (XX) | XX (XX) |
| **Compound sodium lactate solution** |  |  |  |
| N | XX | XX | XX |
| Mean (SD) | XX (XX) | XX (XX) | XX (XX) |
| **Plasma-lyte 148** |  |  |  |
| N | XX | XX | XX |
| Mean (SD) | XX (XX) | XX (XX) | XX (XX) |
| **5% Dextrose** |  |  |  |
| N | XX | XX | XX |
| Mean (SD) | XX (XX) | XX (XX) | XX (XX) |
| **Other dextrose containing fluids** |  |  |  |
| N | XX | XX | XX |
| Mean (SD) | XX (XX) | XX (XX) | XX (XX) |
| **8.4% Sodium bicarbonate** |  |  |  |
| N | XX | XX | XX |
| Mean (SD) | XX (XX) | XX (XX) | XX (XX) |
| **4% Albumin** |  |  |  |
| N | XX | XX | XX |
| Mean (SD) | XX (XX) | XX (XX) | XX (XX) |
| **20% Albumin** |  |  |  |
| N | XX | XX | XX |
| Mean (SD) | XX (XX) | XX (XX) | XX (XX) |
| **Other fluids** |  |  |  |
| N | XX | XX | XX |
| Mean (SD) | XX (XX) | XX (XX) | XX (XX) |
| **Electrolytes replaced up to 72 hours after inclusion in study** |  |  |  |
| **Potassium replacement** |  |  |  |
| N | XX | XX | XX |
| Mean (SD) | XX (XX) | XX (XX) | XX (XX) |
| **Magnesium replacement** |  |  |  |
| N | XX | XX | XX |
| Mean (SD) | XX (XX) | XX (XX) | XX (XX) |
| **Phosphate replacement** |  |  |  |
| N | XX | XX | XX |
| Mean (SD) | XX (XX) | XX (XX) | XX (XX) |

#### 5.1.3

**Table 3.** Patient outcomes.

| Outcome | Raw estimates |  |  | Model estimates |  |
| --- | --- | --- | --- | --- | --- |
|  | Plasma-Lyte® 148<br>(N = ) | 0.9% saline<br>(N = ) | Total<br>(N = ) | MD, OR or HR (95% CI) | P-value |
| **Primary outcome: Days alive and out of hospital up to 28 days** |  |  |  |  |  |
| Unadjusted analysis | N Mean (SD) | N Mean (SD) | N Mean (SD) | xx.x (xx.x to xx.x) | 0.xxx |
| Adjusted analysis | N Mean (SD) | N Mean (SD) | N Mean (SD) | xx.x (xx.x to xx.x) | 0.xxx |
| Multiple imputations | N Mean (SD) | N Mean (SD) | N Mean (SD) | xx.x (xx.x to xx.x) | 0.xxx |
| **Secondary outcomes** |  |  |  |  |  |
| *Continuous outcomes* |  |  |  |  |  |
| Days alive and out of ICU up to 28 days | N Mean (SD) | N Mean (SD) | N Mean (SD) | xx.x (xx.x to xx.x) |  |
| Number of ICU readmissions up to 28 days after study enrolment | N Mean (SD) | N Mean (SD) | N Mean (SD) | xx.x (xx.x to xx.x) |  |
| Number of hospital readmissions up to 28 days after study enrolment | N Mean (SD) | N Mean (SD) | N Mean (SD) | xx.x (xx.x to xx.x) |  |
| Number of ED readmissions up to 28 days after study enrolment | N Mean (SD) | N Mean (SD) | N Mean (SD) | xx.x (xx.x to xx.x) |  |
| Number of GP visits up to 28 days after study enrolment | N Mean (SD) | N Mean (SD) | N Mean (SD) | xx.x (xx.x to xx.x) |  |
| Number of episodes of post-randomisation decrease in GCS by more than 2 in the first 24 hours | N Mean (SD) | N Mean (SD) | N Mean (SD) | xx.x (xx.x to xx.x) |  |
| Cumulative insulin dosage in the first 48 hours | N Mean (SD) | N Mean (SD) | N Mean (SD) | xx.x (xx.x to xx.x) |  |
| Duration of intravenous insulin infusion in the ICU | N Mean (SD) | N Mean (SD) | N Mean (SD) | xx.x (xx.x to xx.x) |  |
| Cumulative potassium replacement in the first 48 hours | N Mean (SD) | N Mean (SD) | N Mean (SD) | xx.x (xx.x to xx.x) |  |
| *Categorical outcomes* |  |  |  |  |  |
| Acute Kidney Injury using KDIGO criteria |  |  |  |  |  |
| *Stage 1* | xx/xxx (xx.x%) | xx/xxx (xx.x%) | xx/xxx (xx.x%) | Reference |  |
| *Stage 2* | xx/xxx (xx.x%) | xx/xxx (xx.x%) | xx/xxx (xx.x%) | xx.x (xx.x to xx.x) |  |
| *Stage 3* | xx/xxx (xx.x%) | xx/xxx (xx.x%) | xx/xxx (xx.x%) | xx.x (xx.x to xx.x) |  |
| Hypokalaemia | xx/xxx (xx.x%) | xx/xxx (xx.x%) | xx/xxx (xx.x%) | xx.x (xx.x to xx.x) |  |
| *Survival outcomes* |  |  |  |  |  |
| Time to hospital discharge | N event / median survival time | N event / median survival time | N event / median survival time | xx.x (xx.x to xx.x) |  |
| Time to ICU discharge | N event / median survival time | N event / median survival time | N event / median survival time | xx.x (xx.x to xx.x) |  |
| Time to resolution of ketosis | N event / median survival time | N event / median survival time | N event / median survival time | xx.x (xx.x to xx.x) |  |

#### 5.1.4

**Table 4.** Summary of adverse events.

| Adverse Events | Plasma-Lyte® |  | P-value |
| --- | --- | --- | --- |
|  | 148<br>(N = ) | 0.9% saline<br>(N = ) |  |
| Adverse Drug Reactions | nEVT nPT(xx.x%) | nEVT nPT(xx.x%) | 0.xx |
| Serious Adverse Drug Reactions | nEVT nPT(xx.x%) | nEVT nPT(xx.x%) | 0.xx |
| Suspected Unexpected Serious Adverse Reactions | nEVT nPT(xx.x%) | nEVT nPT(xx.x%) | 0.xx |
| Serious adverse events | nEVT nPT(xx.x%) | nEVT nPT(xx.x%) | 0.xx |
| Serious events |  |  |  |
| Resulted in death | nEVT nPT(xx.x%) | nEVT nPT(xx.x%) |  |
| Life threatening | nEVT nPT(xx.x%) | nEVT nPT(xx.x%) |  |
| Requires prolonged hospitalisation | nEVT nPT(xx.x%) | nEVT nPT(xx.x%) |  |
| Results in persistent or severe disability/incapacity | nEVT nPT(xx.x%) | nEVT nPT(xx.x%) |  |
| Results in congenital anomaly/birth defect | nEVT nPT(xx.x%) | nEVT nPT(xx.x%) |  |
| Medically significant | nEVT nPT(xx.x%) | nEVT nPT(xx.x%) |  |
| Relationship to study treatment |  |  |  |
| Not related | nEVT nPT(xx.x%) | nEVT nPT(xx.x%) |  |
| Unlikely related | nEVT nPT(xx.x%) | nEVT nPT(xx.x%) |  |
| Possibly related | nEVT nPT(xx.x%) | nEVT nPT(xx.x%) |  |
| Probably related | nEVT nPT(xx.x%) | nEVT nPT(xx.x%) |  |
| Definitely related | nEVT nPT(xx.x%) | nEVT nPT(xx.x%) |  |

#### 5.1.5

**Table 5.** Adverse events of special interest (AESI)

| Event | Plasma-Lyte®<br>148<br>(N = XXX) | 0.9% saline<br>(N = XXX) | Total<br>(N = XXX) |
| --- | --- | --- | --- |
| Number of Hypoglycaemia episodes | XX (XX%) | XX (XX%) | XX (XX%) |
| Number of Hypophosphataemia episodes | XX (XX%) | XX (XX%) | XX (XX%) |
| Number of Hypernatraemia episodes | XX (XX%) | XX (XX%) | XX (XX%) |
| Number of Episodes of Glasgow Coma<br>Score decrease by 2 or more points | XX (XX%) | XX (XX%) | XX (XX%) |

### 5.2 Figures

#### 5.2.1

**Figure 1.**
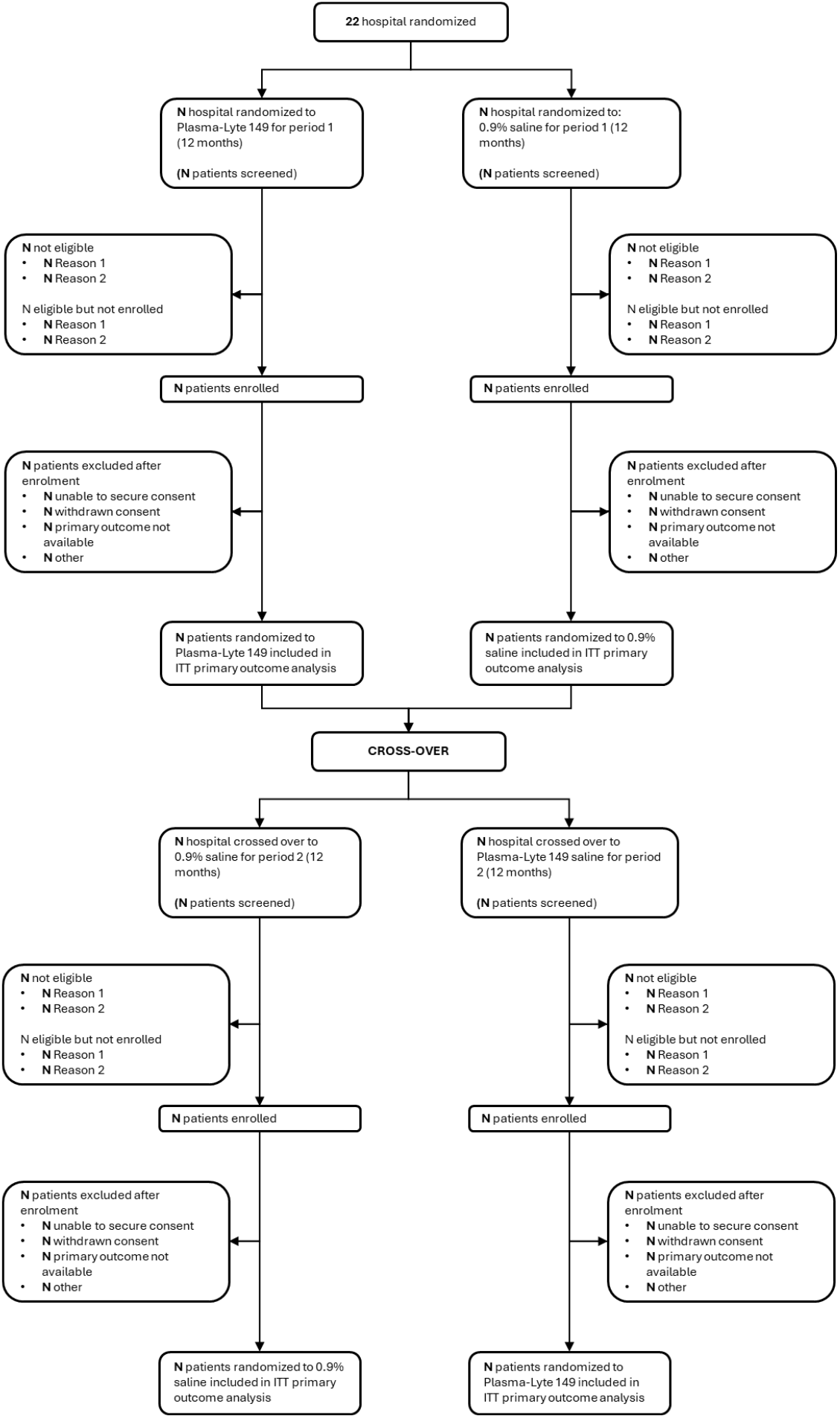
CONSORT diagram.

#### 5.2.2

**Figure 2.** Panel Boxplot of biochemical data over time (can be 1 panel of 8 or 2 panels of 4) Plots will consist of means and 95% confidence bands by treatment arm at baseline, 6, 12, 24, 36, 48 and 72 hours after enrolment for the following biochemical data:

- Sodium
- Chloride
- Potassium
- pH
- Standard base excess
- Anion gap
- Glucose
- Ketones

#### 5.2.3

**Figure 3.** Cumulative incidence function plots. Plots will consist of cumulative incidence up to 28 days by treatment arm for the following outcome data:

- Discharge from critical care area
- Discharge from hospital Programming note: add number at risk every 7 days, median, quartiles, hazard ratio, 95% CI and p-value from the Cox model.

#### 5.2.4

**Figure 4.** Forest plot for subgroup analysis of days alive and out of hospital.

## 6 Supplementary tables and figures

### 6.1 Tables

#### 6.1.1

**Table 1.** Process Measures.

| Measure | Plasma-Lyte®<br>148<br>(N = XXX) | 0.9% saline<br>(N = XXX) | Total<br>(N = XXX) |
| --- | --- | --- | --- |
| **Serum acetoacetate at 12 hours** |  |  |  |
| N | XX | XX | XX |
| Mean (SD) | XX (XX) | XX (XX) | XX (XX) |
| Median (Q1 – Q3) | XX (XX – XX) | XX (XX – XX) | XX (XX – XX) |
| Min - Max | XX - XX | XX - XX | XX - XX |
| **Serum acetoacetate at 24 hours** |  |  |  |
| N | XX | XX | XX |
| Mean (SD) | XX (XX) | XX (XX) | XX (XX) |
| Median (Q1 – Q3) | XX (XX – XX) | XX (XX – XX) | XX (XX – XX) |
| Min - Max | XX - XX | XX - XX | XX - XX |
| **Serum acetoacetate at 48 hours** |  |  |  |
| N | XX | XX | XX |
| Mean (SD) | XX (XX) | XX (XX) | XX (XX) |
| Median (Q1 – Q3) | XX (XX – XX) | XX (XX – XX) | XX (XX – XX) |
| Min - Max | XX - XX | XX - XX | XX - XX |
| **Serum beta-hydroxybutyrate at 12 hours** |  |  |  |
| N | XX | XX | XX |
| Mean (SD) | XX (XX) | XX (XX) | XX (XX) |
| Median (Q1 – Q3) | XX (XX – XX) | XX (XX – XX) | XX (XX – XX) |
| Min – Max | XX - XX | XX - XX | XX - XX |
| **Serum beta-hydroxybutyrate at 24 hours** |  |  |  |
| N | XX | XX | XX |
| Mean (SD) | XX (XX) | XX (XX) | XX (XX) |
| Median (Q1 – Q3) | XX (XX – XX) | XX (XX – XX) | XX (XX – XX) |
| Min - Max | XX - XX | XX - XX | XX - XX |
| **Serum beta-hydroxybutyrate at 48 hours** |  |  |  |
| N | XX | XX | XX |
| Mean (SD) | XX (XX) | XX (XX) | XX (XX) |
| Median (Q1 – Q3) | XX (XX – XX) | XX (XX – XX) | XX (XX – XX) |
| Min - Max | XX - XX | XX - XX | XX - XX |
| **Serum base excess at 6 hours** |  |  |  |
| N | XX | XX | XX |
| Mean (SD) | XX (XX) | XX (XX) | XX (XX) |
| Median (Q1 – Q3) | XX (XX – XX) | XX (XX – XX) | XX (XX – XX) |
| Min - Max | XX - XX | XX - XX | XX - XX |
| **Serum base excess at 12 hours** |  |  |  |
| N | XX | XX | XX |
| Mean (SD) | XX (XX) | XX (XX) | XX (XX) |
| Median (Q1 – Q3) | XX (XX – XX) | XX (XX – XX) | XX (XX – XX) |
| Min - Max | XX - XX | XX - XX | XX - XX |
| **Serum base excess at 24 hours** |  |  |  |
| N | XX | XX | XX |
| Mean (SD) | XX (XX) | XX (XX) | XX (XX) |
| Median (Q1 – Q3) | XX (XX – XX) | XX (XX – XX) | XX (XX – XX) |
| Min - Max | XX - XX | XX - XX | XX - XX |
| **Serum sodium at 24 hours** |  |  |  |
| N | XX | XX | XX |
| Mean (SD) | XX (XX) | XX (XX) | XX (XX) |
| Median (Q1 – Q3) | XX (XX – XX) | XX (XX – XX) | XX (XX – XX) |
| **Serum potassium at 24 hours** |  |  |  |
| N | XX | XX | XX |
| Mean (SD) | XX (XX) | XX (XX) | XX (XX) |
| Median (Q1 – Q3) | XX (XX – XX) | XX (XX – XX) | XX (XX – XX) |
| **Serum chloride at 24 hours** |  |  |  |
| N | XX | XX | XX |
| Mean (SD) | XX (XX) | XX (XX) | XX (XX) |
| Median (Q1 – Q3) | XX (XX – XX) | XX (XX – XX) | XX (XX – XX) |

#### 6.1.3

**Table 2.** Euro-QOL-5D-5L at 28 days.

|  | Plasma-Lyte® 148<br>(N = XXX) | 0.9% saline<br>(N = XXX) | Total<br>(N = XXX) |
| --- | --- | --- | --- |
| **Mobility** |  |  |  |
| I have no problems in walking | xx/xxx (xx.x%) | xx/xxx (xx.x%) | xx/xxx (xx.x%) |
| I have slight problems in walking | xx/xxx (xx.x%) | xx/xxx (xx.x%) | xx/xxx (xx.x%) |
| I have moderate problems in | xx/xxx (xx.x%) | xx/xxx (xx.x%) | xx/xxx (xx.x%) |
| I have severe problems walking | xx/xxx (xx.x%) | xx/xxx (xx.x%) | xx/xxx (xx.x%) |
| I am unable to walk about | xx/xxx (xx.x%) | xx/xxx (xx.x%) | xx/xxx (xx.x%) |
| **Self-care** |  |  |  |
| I have no problems washing or | xx/xxx (xx.x%) | xx/xxx (xx.x%) | xx/xxx (xx.x%) |
| I have slight problems washing or | xx/xxx (xx.x%) | xx/xxx (xx.x%) | xx/xxx (xx.x%) |
| I have moderate problems | xx/xxx (xx.x%) | xx/xxx (xx.x%) | xx/xxx (xx.x%) |
| I have severe problems washing or | xx/xxx (xx.x%) | xx/xxx (xx.x%) | xx/xxx (xx.x%) |
| I am unable to wash or dress | xx/xxx (xx.x%) | xx/xxx (xx.x%) | xx/xxx (xx.x%) |
| **Usual activities** |  |  |  |
| I have no problems doing my usual | xx/xxx (xx.x%) | xx/xxx (xx.x%) | xx/xxx (xx.x%) |
| I have slight problems doing my | xx/xxx (xx.x%) | xx/xxx (xx.x%) | xx/xxx (xx.x%) |
| I have moderate problems doing | xx/xxx (xx.x%) | xx/xxx (xx.x%) | xx/xxx (xx.x%) |
| I have severe problems doing my | xx/xxx (xx.x%) | xx/xxx (xx.x%) | xx/xxx (xx.x%) |
| I am unable to do my usual | xx/xxx (xx.x%) | xx/xxx (xx.x%) | xx/xxx (xx.x%) |
| **Pain / discomfort** |  |  |  |
| I have no pain or discomfort | xx/xxx (xx.x%) | xx/xxx (xx.x%) | xx/xxx (xx.x%) |
| I have slight pain or discomfort | xx/xxx (xx.x%) | xx/xxx (xx.x%) | xx/xxx (xx.x%) |
| I have moderate pain or discomfort | xx/xxx (xx.x%) | xx/xxx (xx.x%) | xx/xxx (xx.x%) |
| I have severe pain or discomfort | xx/xxx (xx.x%) | xx/xxx (xx.x%) | xx/xxx (xx.x%) |
| I have extreme pain or discomfort | xx/xxx (xx.x%) | xx/xxx (xx.x%) | xx/xxx (xx.x%) |
| **Anxiety / depression** |  |  |  |
| I am not anxious or depressed | xx/xxx (xx.x%) | xx/xxx (xx.x%) | xx/xxx (xx.x%) |
| I am slightly anxious or depressed | xx/xxx (xx.x%) | xx/xxx (xx.x%) | xx/xxx (xx.x%) |
| I am moderately anxious or | xx/xxx (xx.x%) | xx/xxx (xx.x%) | xx/xxx (xx.x%) |
| I am severely anxious or | xx/xxx (xx.x%) | xx/xxx (xx.x%) | xx/xxx (xx.x%) |
| I am extremely anxious or | xx/xxx (xx.x%) | xx/xxx (xx.x%) | xx/xxx (xx.x%) |
| **VAS** |  |  |  |
| N Non-missing | XX | XX | XX |
| Mean (SD) | XX (XX) | XX (XX) | XX (XX) |
| Median (Q1-Q3) | XX (XX-XX) | XX (XX-XX) | XX (XX-XX) |
| Min - Max | XX - XX | XX - XX | XX - XX |
| **EQ-5D utility score using**<br>**Australian population norms** |  |  |  |
| N Non-missing | XX | XX | XX |
| Mean (SD) | XX (XX) | XX (XX) | XX (XX) |
| Median (Q1-Q3) | XX (XX-XX) | XX (XX-XX) | XX (XX-XX) |
| Min - Max | XX - XX | XX - XX | XX - XX |

#### 6.1.3

**Table 3.** Modified fatigue scale.

|  | Plasma-Lyte® 148<br>(N = XXX) | 0.9% saline<br>(N = XXX) | Total<br>(N = XXX) |
| --- | --- | --- | --- |
| **Physical subscale (0-36)** |  |  |  |
| N Non-missing | XX | XX | XX |
| Mean (SD) | XX (XX) | XX (XX) | XX (XX) |
| Median (Q1-Q3) | XX (XX-XX) | XX (XX-XX) | XX (XX-XX) |
| Min - Max | XX - XX | XX - XX | XX - XX |
| **Cognitive subscale (0-40)** |  |  |  |
| N Non-missing | XX | XX | XX |
| Mean (SD) | XX (XX) | XX (XX) | XX (XX) |
| Median (Q1-Q3) | XX (XX-XX) | XX (XX-XX) | XX (XX-XX) |
| Min - Max | XX - XX | XX - XX | XX - XX |
| **Psychosocial subscale (0-8)** |  |  |  |
| N Non-missing | XX | XX | XX |
| Mean (SD) | XX (XX) | XX (XX) | XX (XX) |
| Median (Q1-Q3) | XX (XX-XX) | XX (XX-XX) | XX (XX-XX) |
| Min - Max | XX - XX | XX - XX | XX - XX |
| **Total score (0-84)** |  |  |  |
| N Non-missing | XX | XX | XX |
| Mean (SD) | XX (XX) | XX (XX) | XX (XX) |
| Median (Q1-Q3) | XX (XX-XX) | XX (XX-XX) | XX (XX-XX) |
| Min - Max | XX - XX | XX - XX | XX - XX |

#### 6.1.5

**Table 4.** Invasive interventions.

|  | Plasma-Lyte®<br>148<br>(N = XXX) | 0.9% saline<br>(N = XXX) | Total<br>(N = XXX) |
| --- | --- | --- | --- |
| **Mechanical ventilation use** | XX (XX%) | XX (XX%) | XX (XX%) |
| **Vasoactive infusion use** | XX (XX%) | XX (XX%) | XX (XX%) |
| **Renal replacement therapy use** | XX (XX%) | XX (XX%) | XX (XX%) |
| **Catheter use** |  |  |  |
| Central venous catheter | XX (XX%) | XX (XX%) | XX (XX%) |
| Arterial catheter | XX (XX%) | XX (XX%) | XX (XX%) |

#### 6.1.5

**Table 5.** Protocol deviations.

|  | Plasma-Lyte®<br>148 | 0.9% saline<br>(N = XXX) | Total<br>(N = XXX) |
| --- | --- | --- | --- |
| **Protocol deviation** | XX (XX%) | XX (XX%) | XX (XX%) |
| Enrolment of an ineligible patient | XX (XX%) | XX (XX%) | XX (XX%) |
| Non-study fluid administered |  |  |  |
| Saline | XX (XX%) | XX (XX%) | XX (XX%) |
| Plasma-lyte | XX (XX%) | XX (XX%) | XX (XX%) |
| Hartann's | XX (XX%) | XX (XX%) | XX (XX%) |
| Glucose solution | XX (XX%) | XX (XX%) | XX (XX%) |
| Mixed glucose/saline solution | XX (XX%) | XX (XX%) | XX (XX%) |
| Gelatin based fluid | XX (XX%) | XX (XX%) | XX (XX%) |
| Starch based fluid | XX (XX%) | XX (XX%) | XX (XX%) |
| Other | XX (XX%) | XX (XX%) | XX (XX%) |

### 6.2 Figures

### 6.3 Supplementary figures

#### 6.3.1

**Figure 1.** Cumulative incidence function plots. - Resolution of ketosis
- Return of base excess to >= −3 mEq/L
- Time to ADA criteria (ketones<=0.6 AND (pH>=7.30 OR bicarbonate>=18) for DKA resolution
- Cessation of intravenous insulin infusion Programming note: add number at risk every 7 days, median, quartiles, hazard ratio, 95% CI and P value from the Cox model. Any figures from Figs. 2 and 3 which were not able to be put into the panels

## 7. Additional outputs

### 7.1 Tables

#### 7.1.1

**Table 1.**
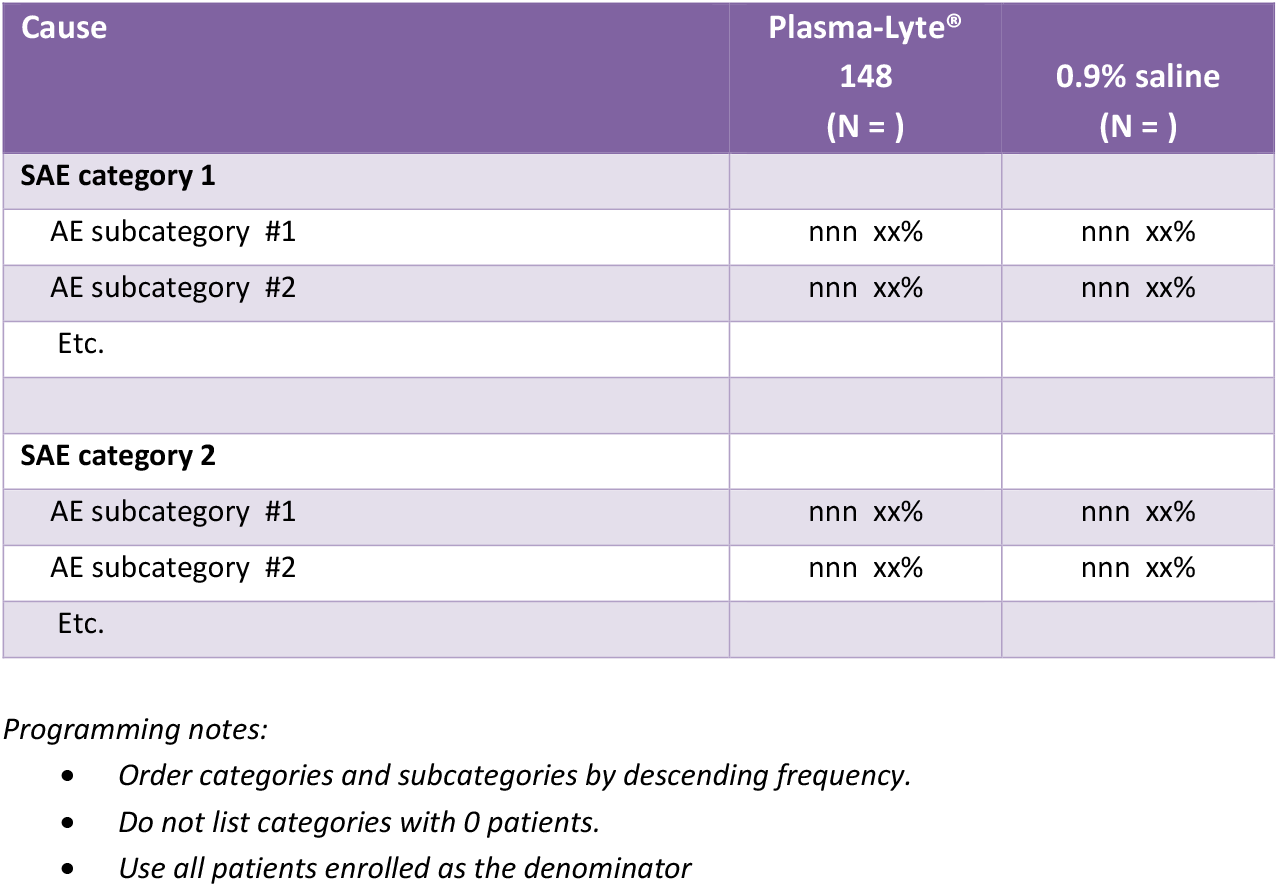
AEs by categories.

### 7.2 Listings

#### 7.2.1

**Listing 1.**
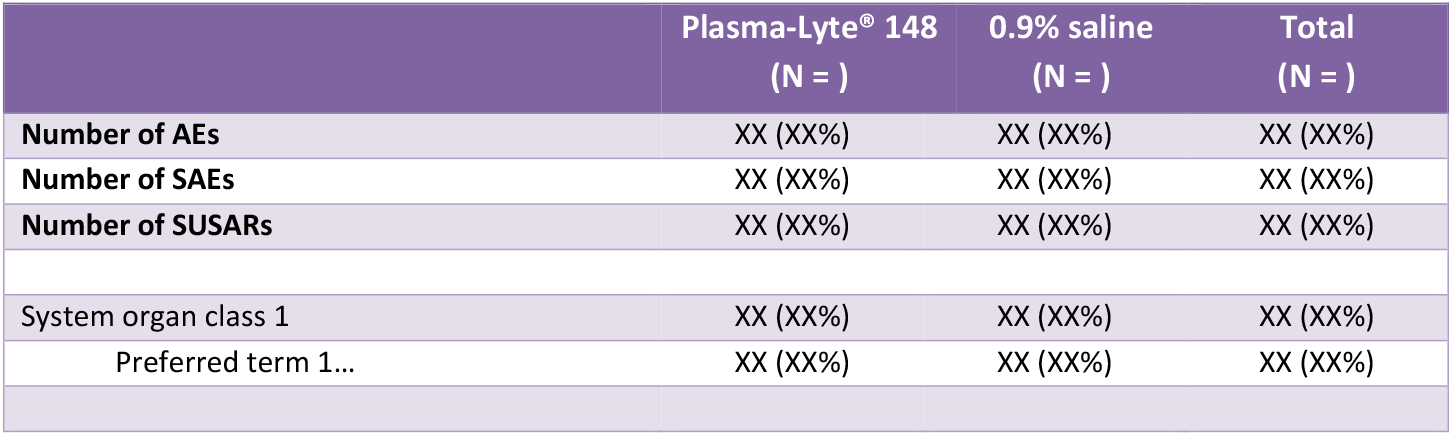
Adverse events.

#### 7.2.2

**Listing 1.** Protocol deviations.

7.2.2 Listing 1. Protocol deviations
| Site ID | Patient ID | Date | Type of deviation | Description | Corrective action taken |
| --- | --- | --- | --- | --- | --- |
| 6 | 6001 | 27-Sep-25 |  |  |  |
|  | 6008 | 20-Sep-25 |  |  |  |
|  | 6052 | 26-Jun-25 |  |  |  |
|  | 6054 | 21-Jun-25 |  |  |  |

## Notes

### Competing Interest Statement

The authors declare the following financial interests/personal relationships which may be considered as potential competing interests: Mahesh Ramanan reports financial support for BEST-DKA
was provided by Medical Research Futures Fund. Gerben Keijzers reports financial support for BEST-DKAwas provided by Emergency Medicine Foundation. Anthony Russell reports financial support for
BEST-DKA was provided by Diabetes Australia. Balasubramanian Venkatesh reports financial support and equipment, drugs, or supplies for BEST-DKA were provided by Baxter Healthcare Ltd. Elif Ekinci reports clinical trial funding from Eli Lilly, Novo Nordisk, Boehringer-Ingelheim, Versanic, Amgen, Novartis and Endogenex. Elif Ekinci sits on advisory boards and given presentations for Bayer,
Eli Lilly, Astra Zeneca, Boehringer and these funds are donated to her institution for diabetes research. The other authors declare that
they have no known competing financial interests or personal relationships that could have appeared to influence the work reported in this paper.

### Clinical Trial

NCT05752279

### Funding Statement

Medical Research Future Fund (MRFF2030670).
EmergencyMedicine Foundation (EMPJ-271R39-2023-KEIJZERS).
Diabetes Australia (Y24M1-RUSA).
Baxter Healthcare.

### Author Declarations

Metro North Human Research Ethics Committee (Royal Brisbane and Women's Hospital) approved the study (approval number: HREC/2022/MNHA/91605).

